# Seasonality and Trends in Stevens-Johnson Syndrome/Toxic Epidermal Necrolysis Before and During the COVID-19 Pandemic: A Pharmacovigilance Study

**DOI:** 10.1101/2025.08.20.25331677

**Authors:** Eric Milan Mukherjee, Dodie Park, Matthew S. Krantz, Cosby A. Stone, Michelle Martin-Pozo, Elizabeth Phillips

## Abstract

**Importance:** **S**easonal variation in adverse drug reactions has clinical and mechanistic implications for understanding disease mechanisms and risk mitigation strategies. Stevens-Johnson Syndrome/Toxic Epidermal Necrolysis (SJS/TEN) is a life-threatening mucocutaneous reaction with high morbidity and mortality, which may have a seasonal component.

**Objective:** To determine whether the reporting of SJS/TEN to the U.S. Food and Drug Administration Adverse Event Reporting System (FAERS) follows a seasonal pattern, incorporating both traditional seasonal analyses and time-series modeling.

**Design:** Cross-sectional, population-based analysis of FAERS reports from January 2010 to December 2019. Seasonal differences were assessed using Kruskal-Wallis tests and Seasonal-Trend Decomposition using Loess (STL). Seasonal autoregressive integrated moving average (SARIMA) models were used to counterfactually forecast SJS/TEN and comparator conditions during the COVID-19 pandemic, assessing changes in reporting.

**Setting:** Population-based analysis of spontaneous adverse event reports submitted to FAERS.

**Participants:** All deduplicated FAERS reports with complete event dates from 2010 to 2019 were included. SJS/TEN cases were identified using standardized MedDRA terms. Comparator analyses of known seasonal conditions – photosensitivity reactions, influenza, and respiratory syncytial virus (RSV) – served as positive controls.

**Exposures:** Drug exposures as recorded in FAERS.

**Main Outcomes and Measures:** The primary outcome was the monthly and seasonal proportion of unique SJS/TEN reports, normalized using all FAERS reports during a particular interval as the denominator. Seasonality strength was quantified from STL decomposition (range 0-1). SARIMA models were applied to pre-COVID data to counterfactually forecast trends from March 2020 to December 2023. Forecast accuracy was evaluated using mean squared error (MSE), root mean squared error (RMSE), and residual diagnostics.

**Results:** Among 5,900 SJS/TEN cases reported from 2010-2019, no significant monthly or seasonal variation was detected (p > 0.05), and seasonality strength was low (0.163). Positive controls (influenza, RSV, photosensitivity) showed expected strong seasonality. SARIMA forecasts indicated a mild increase in SJS/TEN reporting during the pandemic, compared to its previous declining trend. Influenza and RSV dropped below predictions during the pandemic, while photosensitivity remained relatively consistent.

**Conclusions and Relevance:** SJS/TEN reporting to FAERS does not exhibit apparent seasonality, in contrast to positive controls. Time-series modeling confirmed these findings and highlighted the relative stability of SJS/TEN reporting during the pandemic compared to respiratory viruses.

## Introduction

Stevens-Johnson Syndrome (SJS) and Toxic Epidermal Necrolysis (TEN) are rare, life-threatening adverse reactions characterized by mucocutaneous blistering.^1^ SJS/TEN is linked to particular medications, including allopurinol, antibiotics, and anticonvulsants.^2^ The mechanisms by which drugs trigger severe cutaneous adverse reactions (SCARs) involve complex gene-environment interactions, particularly human leukocyte antigen (HLA) alleles that confer susceptibility in specific populations.^3^ Despite these insights, it remains unclear whether environmental factors such as seasonality may influence the risk of SJS/TEN via infectious triggers or prescribing patterns.

Seasonal variation has been described for several drug-induced conditions, raising the possibility that environmental or temporal factors may influence the risk of SJS/TEN.^4–6^ However, studies evaluating seasonality in SJS/TEN have yielded inconsistent results.^7,8^ These studies were often constrained by small sample sizes, limited geographical scope, and reliance on hospital-based cohorts. Furthermore, it is unknown how the COVID-19 pandemic has affected the incidence and reporting of SJS/TEN, and there is some suggestion that COVID infection or vaccination can increase risk.^9–11^ The pandemic also saw a decrease in influenza, respiratory syncytial virus (RSV), and *Mycoplasma pneumoniae* (a known cause of an SJS-like erythema multiforme), though *Mycoplasma* had a subsequent post-pandemic resurgence.^12–14^

To address this gap, we investigated potential seasonal patterns in SJS/TEN reporting using the U.S. Food and Drug Administration’s Adverse Event Reporting System (FAERS), a global pharmacovigilance database frequently used to study rare reactions, such as SJS/TEN.^15,16^ We combined traditional seasonality testing with time-series methods to quantify seasonality strength and forecast reporting trends. As positive controls, we assessed influenza, respiratory syncytial virus (RSV), and photosensitivity reactions, all of which have well-characterized seasonal patterns.

## Methods

All data manipulation and analysis were conducted in RStudio 2024.12.0 Build 467 with R version 4.4.2, with the assistance of GPT4. Further methodological details are available in the **eAppendix.**

### Study Population

A previously sanitized, deduplicated version of FAERS was pared down to reports whose event date (event_dt) contained both a month and a year and fell between January 2010 and December 2019.^15^ Cases were subsequently filtered to those reported from the Northern Hemisphere (**eAppendix 1**). Influenza, RSV, and photosensitivity cases were selected from the FAERS’ indication table, while additional photosensitivity cases and all SJS/TEN cases were chosen from the outcome table. For SJS/TEN cases, primary suspect drugs were extracted for subgroup analyses and were considered causative for the sake of this analysis. By contrast, a patient/case was considered exposed to a particular drug if it appeared in the patient’s drug list (in any role). Standard MedDRA terms used for this selection are found in **eAppendix 2**. For time-series analyses, FAERS database was filtered to events between January 2010 and December 2023 (rather than ending at 2019) for both model fitting and counterfactual forecasting.

### Statistical Analyses

For each condition and drug, we calculated monthly and seasonal proportions relative to the total reports submitted per month to accounting for underlying fluctuations in reporting. Seasonal periods were defined using Northern Hemisphere conventions: Winter (December-February), Spring (March-May), Summer (June-August), and Fall (September-November).^8^ Differences in reporting proportions across months and seasons were assessed using Kruskal-Wallis tests, with p-values adjusted for multiple testing using the Benjamini-Hochberg procedure. To further refine our analysis and mitigate confounding from broader reporting trends or seasonal biases in drug exposures, we implemented additional normalizations:

1. Normalization by drug mentions – proportion of SJS/TEN cases per month relative to the number of FAERS reports mentioning the same drug, to account for seasonality in drug exposure/prescribing.
2. Normalization by total SJS/TEN reports – proportion of SJS/TEN cases attributed to each drug per month, normalized by the total number of SJS/TEN reports, to isolate drug-specific contributions to SJS/TEN.

These parallel approaches were designed to mitigate confounding from seasonality in drug prescribing, exposure patterns, or reporting behaviors. For culprits of interest, we also calculated the proportion of patients in FAERS exposed to the drug, normalized by the total number of FAERS reports, to analyze seasonality in drug exposure. For each normalized metric, we applied Kruskal-Wallis tests for both month and season, with Benjamini-Hochberg correction applied within each comparison set.

To quantify the strength of seasonality in each outcome, we applied Seasonal-Trend Decomposition using Loess (STL), which additively partitions time series data into long-term trend, seasonal, and residual components.^17,18^ Seasonality strength was defined as the proportion of variance explained by the seasonal component, scaled between 0 and 1, with 0.3 and 0.6 serving as thresholds for moderate and strong seasonality (**eAppendix 3**).

We implemented Seasonal Autoregressive Integrated Moving Average (SARIMA) models to model temporal dynamics and counterfactually forecast trends during the COVID-19 pandemic. ARIMA-based time-series forecasting has been used to analyze responses to the pandemic, including changes in healthcare delivery and suicide rates.^19,20^ The pandemic was marked as starting in March 2020.^21^ SARIMA models were fit to monthly proportions of each outcome from January 2010 through March 2020 and forecasts were generated for April 2020 to December 2023. Forecast performance was assessed using mean squared error (MSE) and root mean squared error (RMSE). Residuals were evaluated using the Ljung-Box test to detect any remaining autocorrelation.

Two sensitivity analyses were conducted to account for potential geographic confounding: (1) restricting the dataset to reports from high-resource Northern Hemisphere countries with robust pharmacovigilance, and (2) restricting to U.S.-originating reports only. Analyses of Southern Hemisphere cases were not conducted due to insufficient sample size (data not shown).

## Results

### Overall SJS/TEN Reporting Is Not Seasonal

A total of 5,900 unique SJS/TEN reports from the Northern Hemisphere between 2010 and 2019 met the inclusion criteria for the primary analysis, resulting in a total of 4,311,170 unique reports. Monthly and seasonal proportions of SJS/TEN reports, calculated as the raw number of reports divided by the total number of FAERS reports submitted during the corresponding period, demonstrated no statistically significant variation. Specifically, Kruskal-Wallis tests for both monthly and seasonal variation yielded non-significant p-values (p > 0.05), indicating the absence of a discernible seasonal or monthly trend in overall SJS/TEN reporting (**Figure 1A and 1B**).

**Figure 1.**
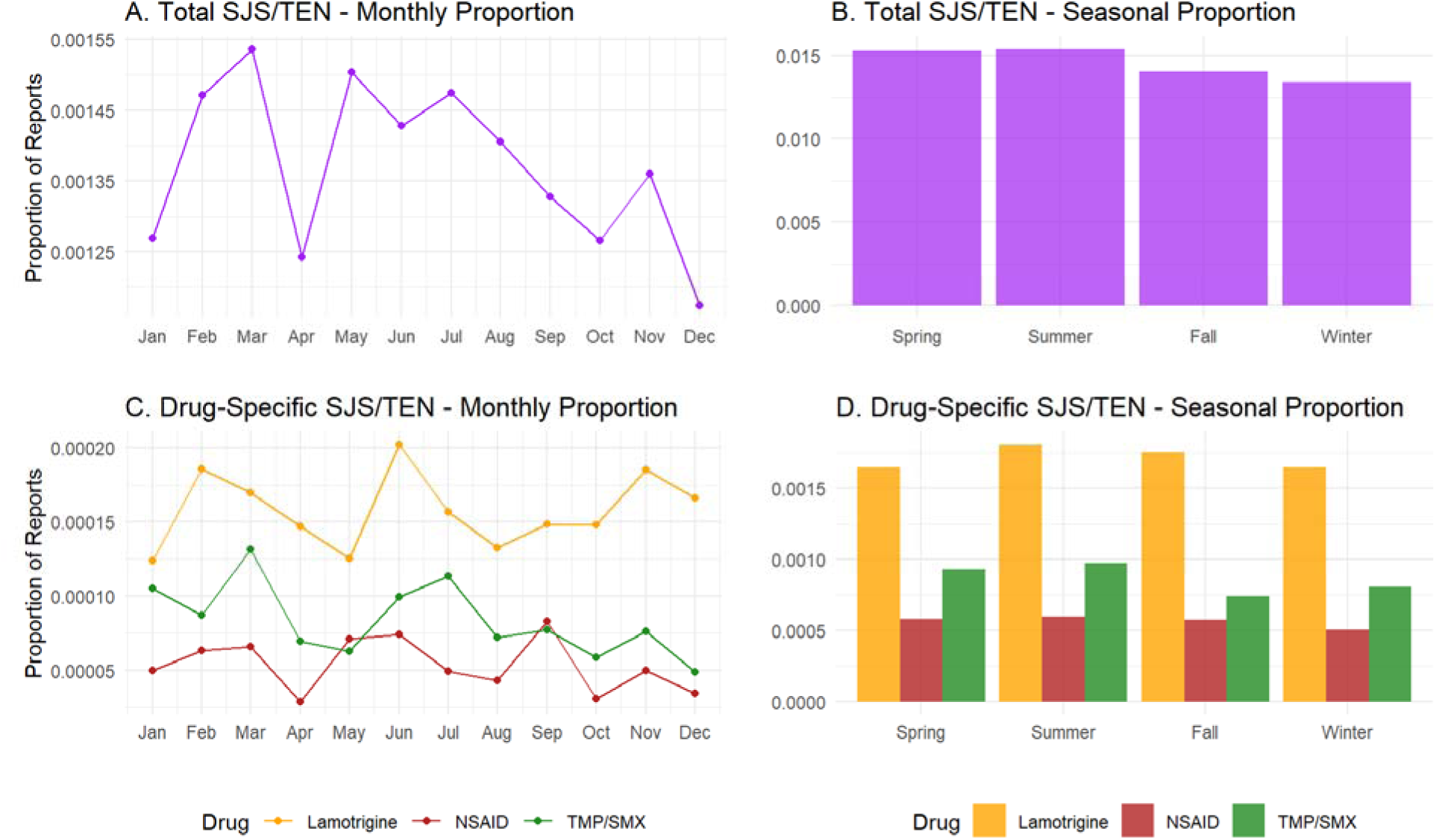
Monthly and Seasonal Patterns in SJS/TEN Reports. Monthly (A, C) and seasonal (B, D) proportions of SJS/TEN reports submitted to FAERS from 2010-2019 are shown for all cases (top panels) and stratified by key drug groups (bottom panels). Proportions represent the number of unique SJS/TEN cases relative to all FAERS reports submitted during each time period. Seasons are defined using Northern Hemisphere convention: Winter (December-February), Spring (March-May), Summer (June-August), and Fall (September-November). Kruskal-Wallis tests for both monthly and seasonal variation were not statistically significant for total SJS/TEN or any individual drug group (all p > 0.05).

We next examined whether cases associated with specific high-risk medications – trimethoprim-sulfamethoxazole (TMP/SMX), over-the-counter nonsteroidal anti-inflammatory drugs (OTCs NSAIDs, namely acetaminophen, naproxen, and ibuprofen), and lamotrigine – exhibited seasonal variation. Among the subset of SJS/TEN reports with these medications as primary suspects, Kruskal-Wallis tests again revealed no statistically significant monthly or seasonal differences (all p > 0.05). These findings were consistent with the overall cohort, suggesting that seasonality does not play a substantial role even when stratified by likely causative agent (**Figure 1C and 1D**).

To further assess underlying seasonal patterns, we decomposed monthly SJS/TEN proportions into long-term trend, seasonal, and residual components using STL decomposition. The results (**Figure 2**) confirmed minimal within-year fluctuations with an overall downward trend in reporting over the 10-year period. The calculated seasonality strength was 0.163. Seasonality strengths for specific primary suspects were uniformly low, with values of 0.102 for TMP/SMX, 0.106 for OTC NSAIDs, and 0.126 for lamotrigine.

**Figure 2.**
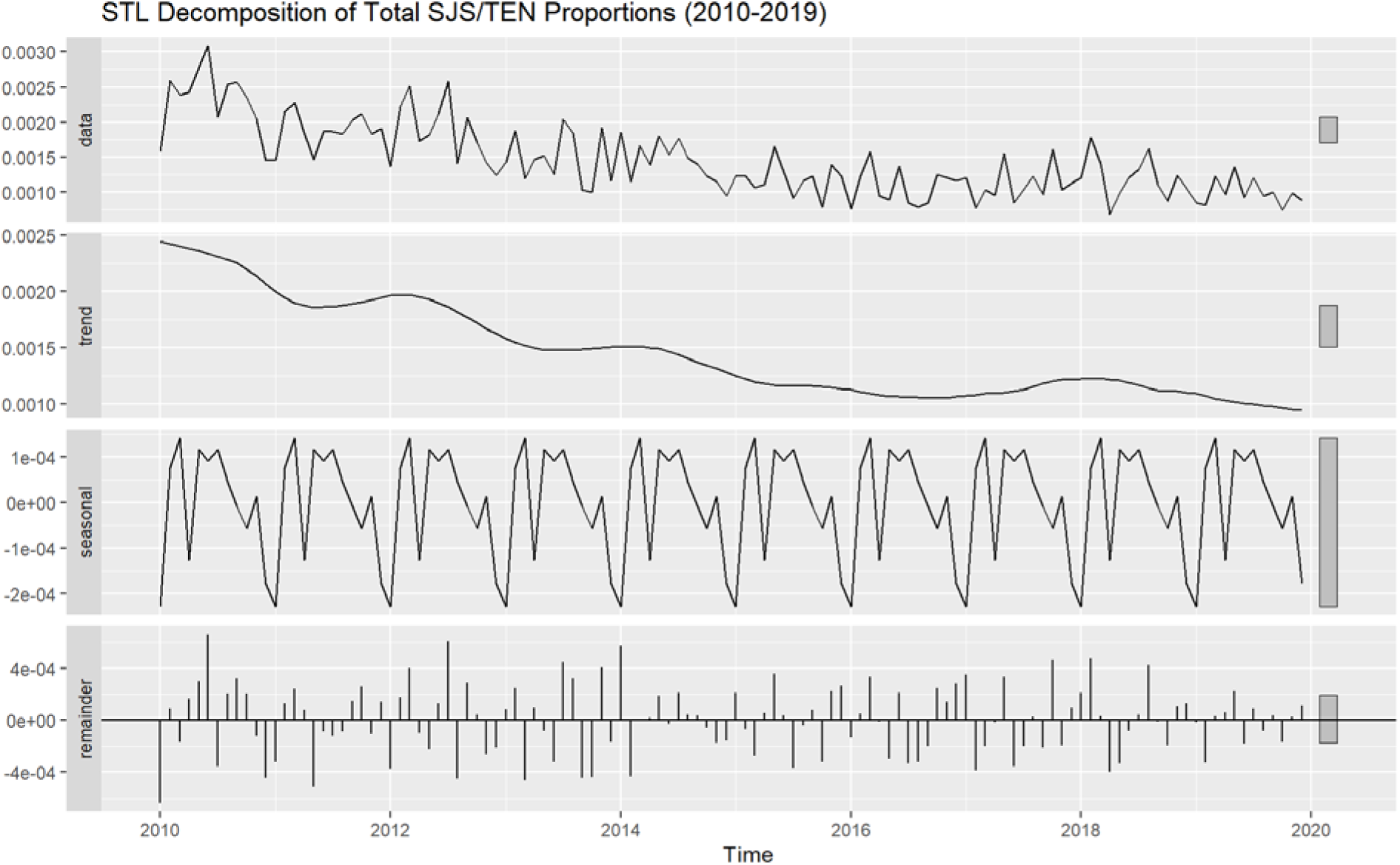
STL Decomposition of SJS/TEN Report Proportions Over Time. The raw time series (top panel) was decomposed into a long-term trend component (second panel), a seasonal component reflecting recurring within-year fluctuations (third panel), and a remainder capturing unexplained variation (bottom panel). Minimal seasonal structure is observed, consistent with low seasonality strength (0.163) calculated from this decomposition.

### Low Seasonality Across Drugs

To validate our methods for detecting seasonal variation, we examined reporting patterns for three positive control indications or outcomes with known seasonal variation: influenza, respiratory syncytial virus (RSV), and photosensitivity. All three demonstrated significant variation by both month and season (**eFigure 1**, Kruskal-Wallis p < 0.001 for each). STL decomposition yielded seasonality strengths of 0.8 for influenza, 0.704 for photosensitivity, and 0.508 for RSV, affirming that our methodology could detect seasonal signals (**eFigures 2-4**).

To further characterize potential seasonal patterns at the drug level, we summarized the seasonality strength, Kruskal-Wallis results, and exposure counts for all drugs serving as primary suspects in at least 50 SJS/TEN cases; seasonality metrics were applied to monthly or seasonal proportions normalized by total FAERS reports (**Table 1**). Consistent with earlier analyses, most drugs exhibited low seasonality strength and non-significant adjusted p-values for both seasonal and monthly variation. Vancomycin exhibits the highest overall seasonality strength among the selected drugs (0.257). Both vancomycin and allopurinol exhibited marginal evidence of seasonal variation, as indicated by Kruskal-Wallis tests; however, this difference became non-significant after adjustment for multiple comparisons. The “No Primary Suspect Listed” category, representing reports without a primary suspect drug, similarly lacked any significant seasonality.

**Table 1.**
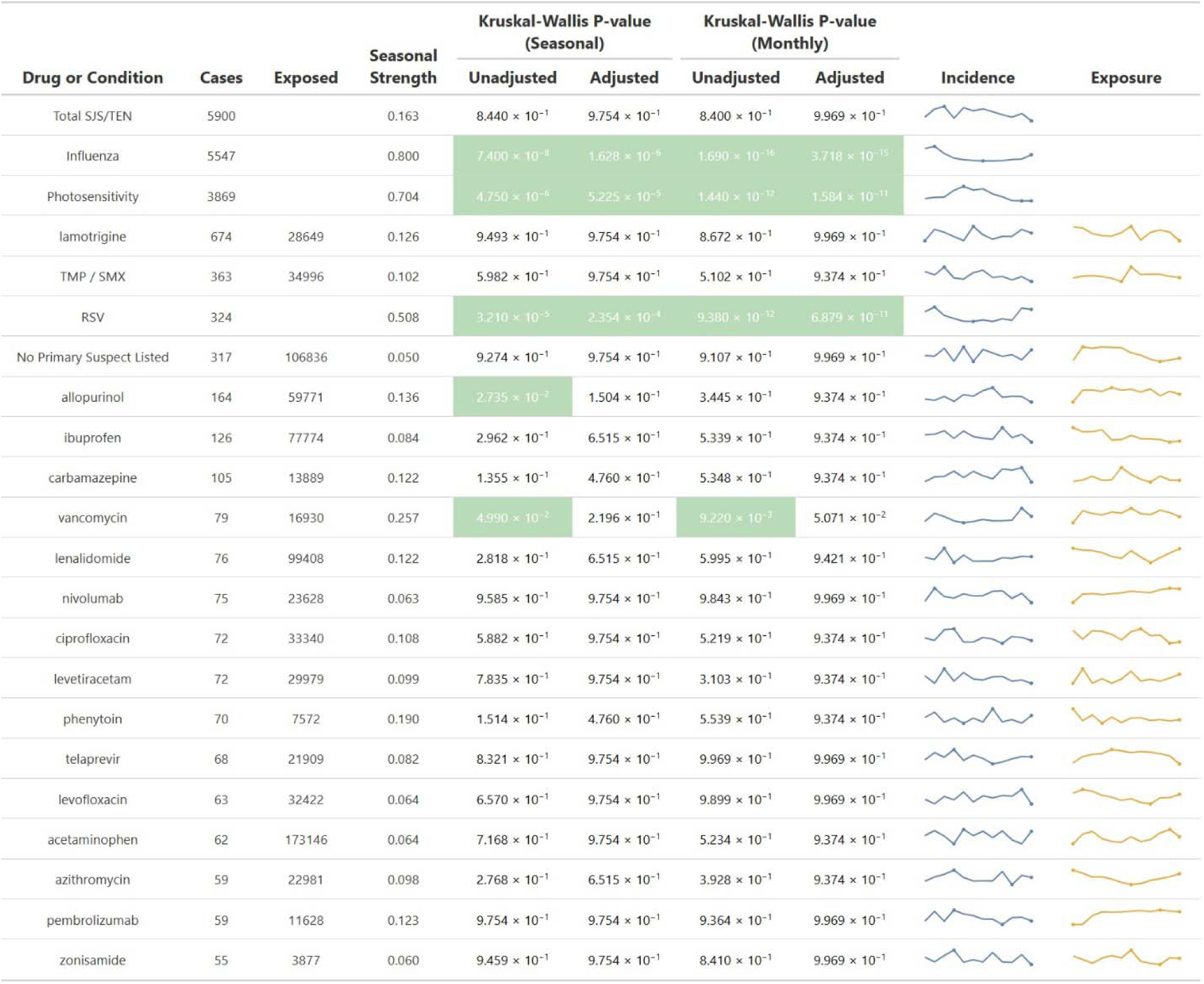
Seasonality Measures – Northern Hemisphere.

In contrast, the positive controls (influenza, photosensitivity, and RSV) remained strongly seasonal with adjusted Kruskal-Wallis p-values < 0.001, and seasonality strengths of 0.8, 0.704, and 0.508, respectively, with the former two demonstrating strong seasonality (>0.6). We also assessed seasonality at the individual drug level using bubble plots to visualize both statistical significance (via –log_10_ adjusted p-values from Kruskal-Wallis tests) and seasonality strength for drugs associated with at least 50 SJS/TEN cases (**Figure 3**). All drugs clustered near the origin, indicating low seasonality strength and minimal evidence of seasonal variation.

**Figure 3.**
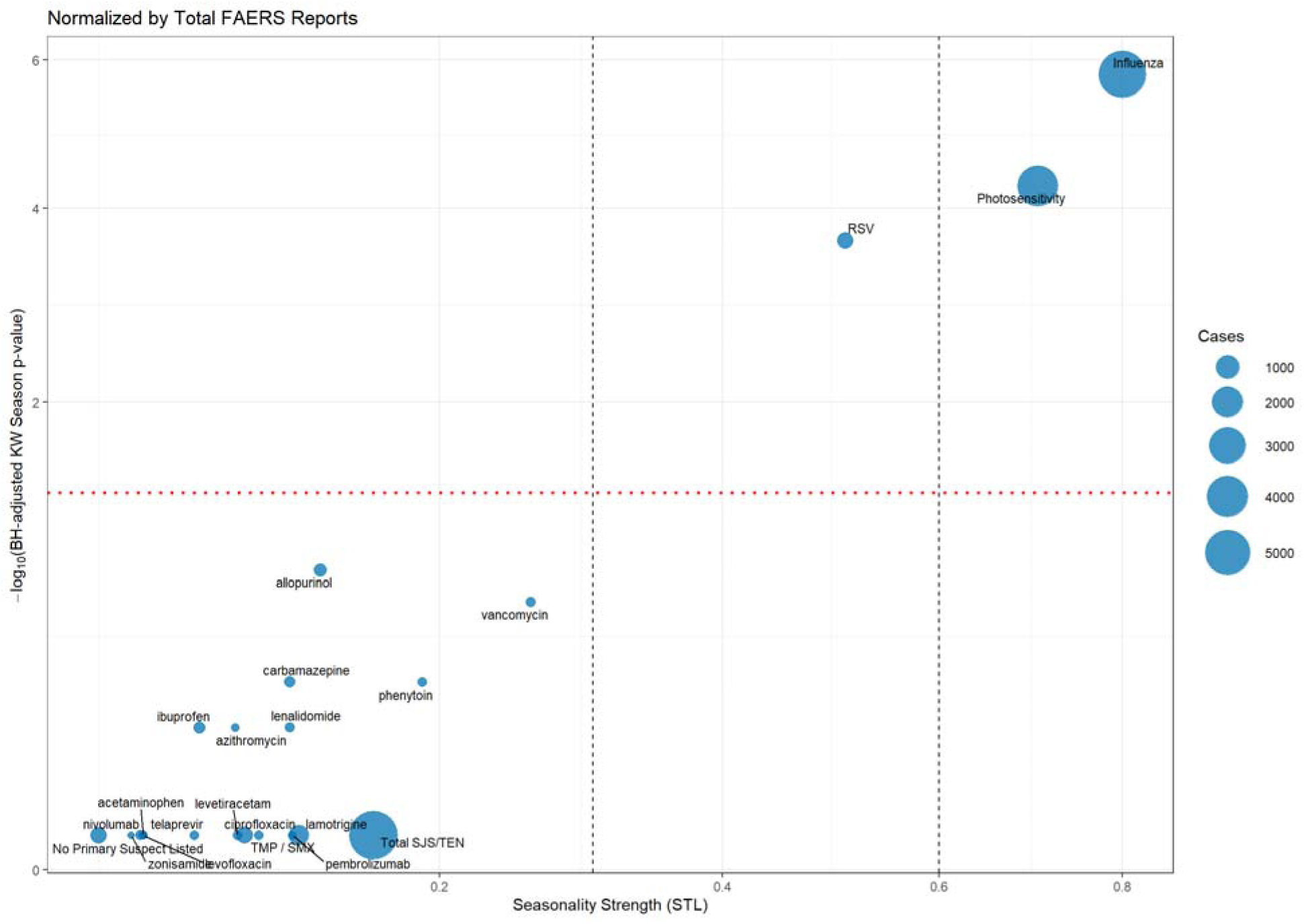
Seasonality Strength and Significance Across Drugs – Northern Hemisphere. Each bubble represents a drug with at least 50 SJS/TEN cases, reported to FAERS from 2010-2019; respiratory viruses (influenza, RSV) and photosensitivity serve as controls. The x-axis shows seasonality strength, calculated from STL decomposition of monthly case proportions (0 = no seasonality, 1 = strong seasonality). The y-axis displays the negative log_10_-transformed Benjamini-Hochberg adjusted p-value from a Kruskal-Wallis test assessing seasonal differences in case proportions. Bubble size reflects the number of cases for each drug. Vertical dashed lines at 0.3 and 0.6 mark thresholds for weak, moderate, and strong seasonality. The horizontal dotted line indicates the significance threshold for Kruskal-Wallis tests (adjusted p = 0.05).

We further explored potential seasonal influences on SJS/TEN reporting by applying multiple normalization strategies designed to subtract broader seasonal effects (**eFigure 5**). Raw reports per drug (panel A) showed a similar lack of seasonality. Panel B illustrates the seasonality of exposure patterns, normalizing patients exposed to each drug per unit time, demonstrating that allopurinol, azithromycin, and ciprofloxacin show some seasonal exposure patterns (with azithromycin prescriptions notably increasing in the winter months, consistent with increased prescribing during respiratory virus season); despite this, their corresponding SJS/TEN cases do not show statistical evidence of seasonality. To determine whether overall SJS/TEN reporting trends affected these measures, we normalized each drug’s reports by the total number of SJS/TEN cases per month (panel C), which also showed a lack of seasonality. Finally, to determine if a seasonal environmental factor could be driving variation, we normalized each drug’s reports by the total number of patients exposed to that drug during the corresponding time period, which normalizes for seasonal variations in exposure/prescribing. This also failed to demonstrate seasonality. Across all normalization methods, SJS/TEN-associated drugs consistently exhibit low seasonality.

To explore potential geographic confounding or artifacts due to regional reporting behavior, we conducted two sensitivity analyses. The first restricted the dataset to reports from a subset of high-resource Northern Hemisphere countries with robust pharmacovigilance infrastructure (**eTable 1**). In contrast, the second included only reports originating from the United States (**eTable 2**). In both cases, the results mirrored the primary analysis: when analyzing the proportion of SJS/TEN reports per drug, normalized to total FAERS reports, no significant monthly or seasonal variation was observed for SJS/TEN, and the strength of seasonality remained low across all subgroups and drug-specific strata. In contrast, positive controls continued to show strong seasonality.

### SARIMA Models Confirm Stability of SJS/TEN Reporting During the COVID-19 Pandemic

To corroborate these findings and evaluate the potential effects of the COVID-19 pandemic on SJS/TEN reporting, we fit SARIMA models to SJS/TEN, influenza, RSV, and photosensitivity data (**eTable 3**). Models were fitted to monthly proportions of each condition relative to all FAERS reports from January 2010 through March 2020. Model residuals passed the Ljung-Box test in all cases (p > 0.05), indicating no significant residual autocorrelation and confirming that models adequately captured underlying temporal patterns.

In contrast, the SARIMA models for comparator conditions demonstrated expected complexity. The model for influenza included multiple seasonal and autoregressive terms capturing annual cycles, while the model for RSV required multiple autoregressive and moving average terms to fit pre-pandemic fluctuations. Photosensitivity exhibited a simpler seasonal model but retained apparent annual periodicity with a single seasonal moving average term (sma1 = −0.879) and a small drift term. In contrast, the SJS/TEN model was notably sparse, consisting of a single moving average term (ma1 = −0.871) and a minimal drift term, reflecting the low seasonality and weak trend in SJS/TEN reporting.

Forecasts from April 2020 to December 2023 revealed distinct patterns across conditions (**Figure 4**). SJS/TEN reporting modestly exceeded forecasted levels, suggesting stable or slightly increased reporting despite pandemic-related disruptions. In contrast, both influenza and RSV reporting sharply diverged from their forecasts, with observed proportions collapsing below predicted values during 2020-2021, consistent with the widespread suppression of respiratory virus transmission during the pandemic.^22^ Notably, influenza exhibited complete abolition of seasonal spikes in the early pandemic years. Conversely, photosensitivity forecasts closely aligned with observed data, demonstrating robust seasonality even during the pandemic.

**Figure 4.**
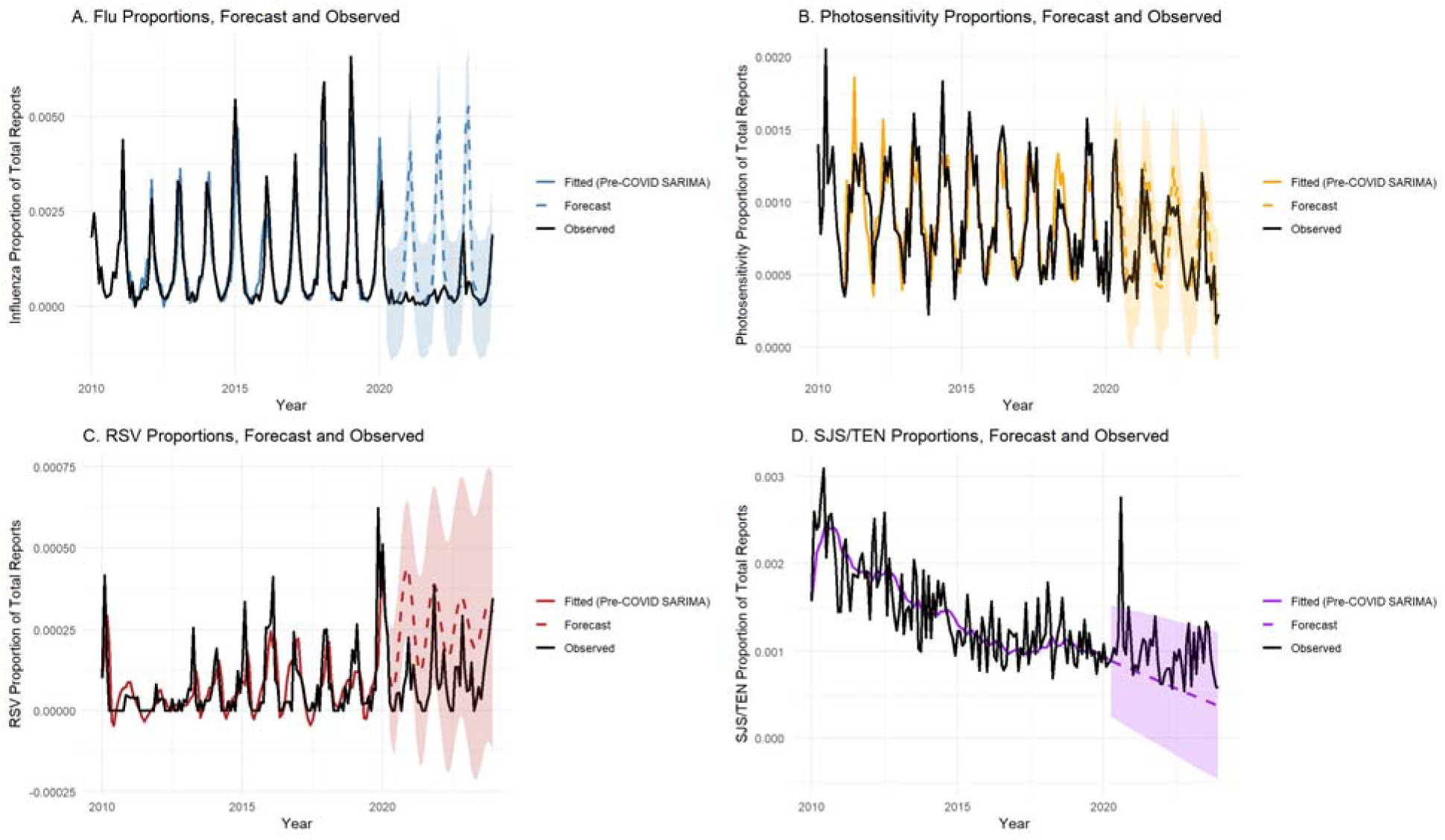
SARIMA Forecasts and Observed Proportions of Adverse Events Before and During the COVID-19 Pandemic. Time series of monthly proportions of adverse event reports submitted to FAERS from 2010 through 2023 for four outcomes: (A) Influenza, (B) Photosensitivity, (C) RSV and (D) SJS/TEN. Solid colored lines represent SARIMA models fitted to pre-pandemic data (2010–March 2020). Dashed lines with shaded areas indicate forecasts and 95% confidence intervals for the post-pandemic period (April 2020–December 2023). Observed data are shown in black. Influenza and RSV reports collapsed below forecasted levels during the pandemic, reflecting disrupted transmission of respiratory viruses. Photosensitivity reports continued to follow forecasted seasonal patterns. In contrast, SJS/TEN reports modestly exceeded forecasts, indicating relative stability or slight proportional increase during the pandemic period despite overall declines in the proportion of total reports over time.

Forecast accuracy metrics further illustrated these dynamics (**eTable 3**). Pre-pandemic MSE and RMSE values were low across all models, indicating a strong fit. However, post-pandemic forecast errors increased substantially for influenza (post-COVID RMSE = 1.68e-03) and RSV (1.83e-04), reflecting the unexpected collapse in reporting. In contrast, SJS/TEN and photosensitivity maintained low post-COVID RMSE and MSE (5.02e-04 and 2.17e-04, respectively), affirming the relative stability of these conditions. The simplicity of the SJS/TEN model, combined with its stable forecasting performance, reinforces the absence of substantial seasonality or pandemic-induced shifts in reporting.

### Erythema Multiforme and Respiratory Mycoplasma do not show Seasonal Trends

We also considered drug-induced erythema multiforme (EM) and EM as indication (aggregated as EM), and respiratory mycoplasma infection (**eFigure 6**). Neither demonstrated seasonality by Kruskal-Wallis or STL decomposition. Corroborating previous reports, the reporting of respiratory mycoplasma sharply increased in 2023.^14^

## Discussion

Many cutaneous eruptions related to drugs and/or viruses have been reported as seasonal, including cytomegalovirus disease in kidney transplant recipients and reactions to iodinated contrast.^5,23,24^ Recently, drug reaction with eosinophilia and systemic symptoms (DRESS) was suggested to have mild seasonality, with fall having slightly increased incidence and mortality.^4^ Previous studies of SJS/TEN seasonality have yielded contradictory results. A single-center study of 13 TMP/SMX-induced-SJS/TEN patients suggested increased incidence in spring, a multicenter European study suggested no seasonality in the proportion of SJS/TEN attributed to TMP/SMX (34 of 377 total SJS/TEN cases), and a study of urban centers in China showed a non-significant increase in the summer (9308 total patients).^7,8,25^

In this pharmacovigilance study of over 5,900 SJS/TEN cases reported to FAERS, we found no evidence of meaningful seasonal variation. Month-to-month and season-to-season changes were statistically nonsignificant, and the strength of seasonality was low across all drugs and subgroups. Time-series decomposition consistently demonstrated minimal variance attributable to seasonal components. These findings were robust to multiple normalization approaches and sensitivity analyses. The stability of SJS/TEN reporting stood in marked contrast to conditions with established seasonality, such as influenza, RSV, and photosensitivity reactions.

The lack of seasonality in SJS/TEN has essential clinical and mechanistic implications. Clinically, it suggests that the risk of SJS/TEN lacks predictable seasonal peaks to inform prevention. Mechanistically, seasonal variation in incidence is often linked to environmental exposures, infection, or prescribing patterns. Our findings reduce the likelihood that such co-factors, such as wintertime infections, sunlight, or variations in drug utilization, substantially modulate SJS/TEN risk. Instead, the data reinforce a model in which idiosyncratic host factors, including genetic predisposition, drug metabolism, and immune responses, predominantly determine susceptibility.^3,26,27^

Time-series forecasting with SARIMA models further underscored the stability of SJS/TEN reporting. Unlike influenza, which collapsed in reporting during the COVID-19 pandemic, SJS/TEN reports slightly exceeded forecasts. This resilience to pandemic-era behavioral and healthcare disruptions supports the notion that SJS/TEN incidence is relatively independent of the seasonal infectious triggers that drive other mucocutaneous conditions. Interestingly, while we have previously shown that the absolute number of global SJS/TEN reports remained stable or slightly increased during this time period^15^, the proportion of SJS/TEN relative to total FAERS reports has declined over time in our data. This likely reflects growth in the overall volume of adverse event reporting, driven by increased pharmacovigilance efforts, heightened reporting of common or non-serious events, and the expanding diversity of therapies on the market. As the denominator of total reports increases, rare events like SJS/TEN may appear proportionally diminished, even when their absolute burden remains unchanged.

There is controversy about a causative link between COVID-19 and/or COVID-19 vaccination and SJS/TEN. A retrospective cohort study found that COVID-19 infection is associated with a 2-fold increased risk of SJS/TEN.^28^ A single-center study in Australia reported a striking seven-fold increase in SJS/TEN incidence during the pandemic, with suggested associations to both COVID-19 infection and vaccination, though its statistical significance is controversial.^10,11,29,30^ Our data contains a single case of SJS/TEN attributed to COVID-19 vaccination (from Italy), and displayed a notable upward deviation in the proportion of SJS/TEN reports during the pandemic years. In addition to the direct effects of COVID-19, this may reflect changes in healthcare engagement, reporting behavior, or heightened surveillance around severe immune-mediated reactions. This divergence between localized incidence data and global pharmacovigilance reporting highlights the complexity of detecting true epidemiological shifts in rare events using spontaneous reporting systems. Further studies integrating population-level incidence data, active surveillance, and robust clinical registries will be essential to clarify whether the pandemic or its countermeasures have influenced the true incidence of SJS/TEN globally.

We explored erythema multiforme (EM) and respiratory *Mycoplasma* infections to address diagnostic overlap with MIRM/RIME and SJS/TEN, which often present similarly but may have distinct triggers. EM demonstrated a borderline seasonal trend with increased reporting in spring and summer (p = 0.053), while *Mycoplasma* showed no seasonal variation (p = 0.802), likely due in part to sparse and inconsistent reporting in pharmacovigilance data. These findings suggest that *Mycoplasma* does not explain EM seasonality and underscore the importance of distinguishing EM from MIRM/RIME in clinical and regulatory settings. However, reporting of respiratory *Mycoplasma* is sparse, likely due to very few adverse events during treatment.

This study has several limitations. FAERS is subject to underreporting, reporting biases, and incomplete information, including imprecise event dates. While we included multiple high-risk drugs, many individual agents were associated with relatively few SJS/TEN cases, limiting our power to detect drug-specific seasonal patterns. We were also unable to formally assess seasonal patterns in the Southern Hemisphere (where reversed seasons could theoretically yield opposite trends), because cases were infrequent. Furthermore, we evaluated seasonality at the reporting level, which may not precisely reflect incidence. Nonetheless, the consistency of findings across drugs, geographic regions, analytic methods, and comparators lends strength to the conclusion that seasonal influences on SJS/TEN are minimal or absent.

In summary, this comprehensive pharmacovigilance study demonstrates that SJS/TEN does not exhibit substantial seasonality, distinguishing it from other drug-related and infectious conditions. These findings enhance our understanding of SJS/TEN pathogenesis and underscore the need for year-round clinical vigilance for this severe reaction. Future studies integrating population-level incidence data, precise onset timing, and genetic risk factors could further elucidate potential environmental contributions to SJS/TEN susceptibility.

## Contributions

EMM conceived the project, wrote the manuscript, and conducted all analyses. DP assisted with analyses and edited the manuscript. MMP, CAS, and MSK assisted with analyses and reviewed the manuscript. EP reviewed the manuscript.

## Supporting information

eAppendix

## Data Availability

All data produced in the present study are available upon reasonable request to the authors

https://github.com/capuhcheeno/

## Acknowledgements

E.J.P. is supported by the following grants from the National Institutes of Health (NIH): NIH U01AI154659, NIH P50GM115305, NIH R01HG010863, NIH R21AI139021, NIH R01AI152183, and NIH 2 D43 TW010559. E.J.P. is also supported by the National Health and Medical Research Council of Australia. E.M.M. is funded by a Vanderbilt University Medical Center internal career development award (Vanderbilt Faculty Research Scholars).

## Conflict of Interest

E.J.P. receives royalties and consulting fees from UpToDate and UpToDate Lexidrug (where she is a Drug Allergy Section Editor and section author) and has received consulting fees from Janssen, Vertex, Verve, Servier, Rapt and Esperion. E.J.P. is co-director of IIID Pty Ltd, which holds a patent for HLA-B*57:01 testing for abacavir hypersensitivity, and E.J.P has a patent pending for detection of HLA-A*32:01 in connection with diagnosing drug reaction with eosinophilia and systemic symptoms to vancomycin. For these patents she does not receive any financial remuneration, and neither is related to the submitted work.

## Statement on the Use of Artificial Intelligence

GPT4o was used to write, optimize, and debug R code and assist with drafting and editing the manuscript for clarity. Suggestions were cross-referenced in literature, package documentation, and resources like stackoverflow.com. All code and generated data were manually reviewed, tested, and verified by the authors. The authors take responsibility for all code generated. GPT4o was also used for idea generation and assistance with clarity in manuscript writing.

---

**eFigure 1.**
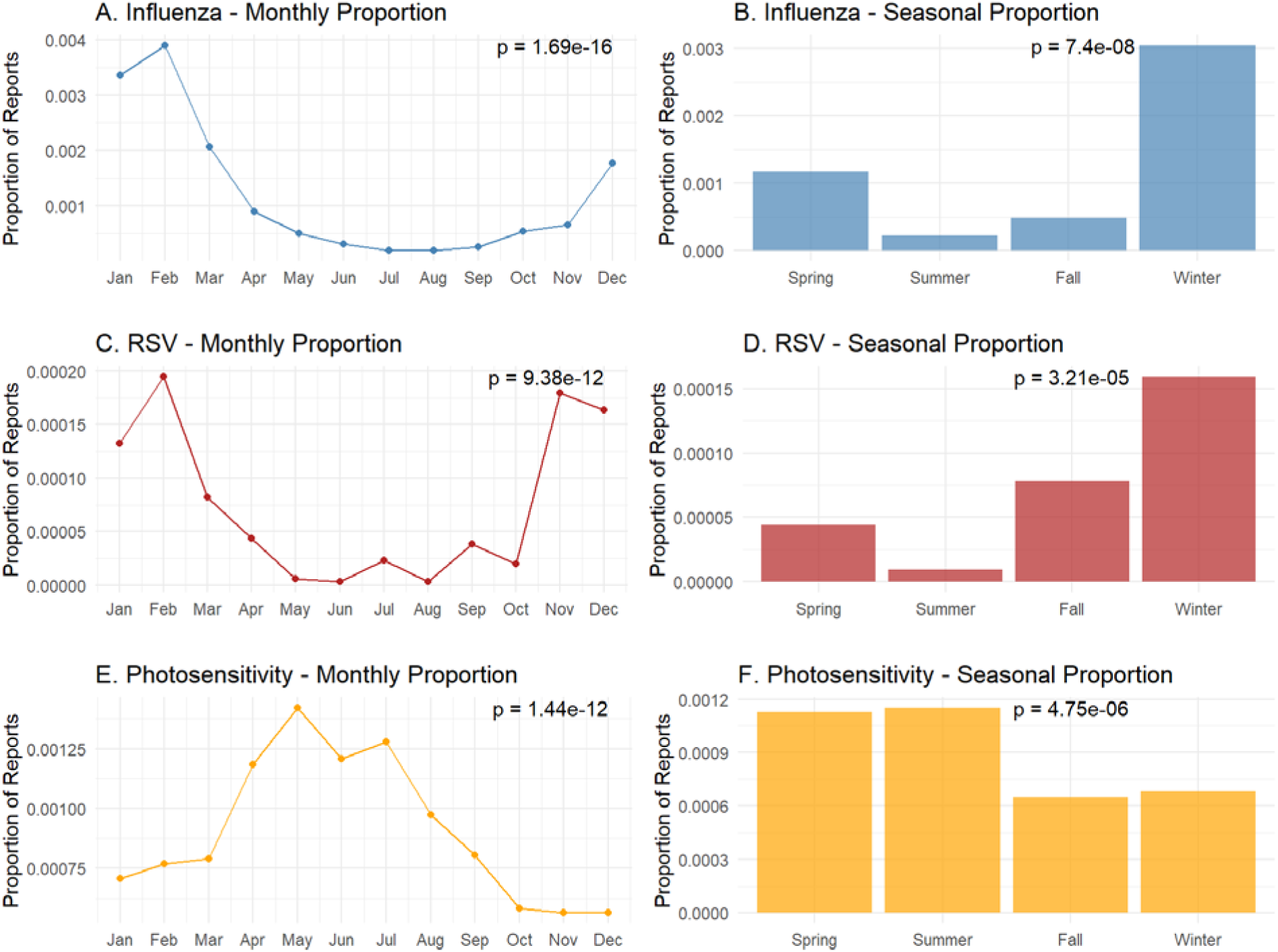
Monthly and Seasonal Patterns in Photosensitivity and Respiratory Virus Reports. Monthly and seasonal proportions of photosensitivity, influenza, and RSV reports submitted to FAERS from 2010-2019 are shown. Proportions represent the number of unique reports for each outcome relative to all FAERS reports submitted during each time period. Seasons are defined using Northern Hemisphere convention: Winter (December-February), Spring (March-May), Summer (June-August), and Fall (September-November). Kruskal-Wallis tests for both monthly and seasonal variation were significant for all three outcomes (p-values shown on each panel).

**eFigure 2.**
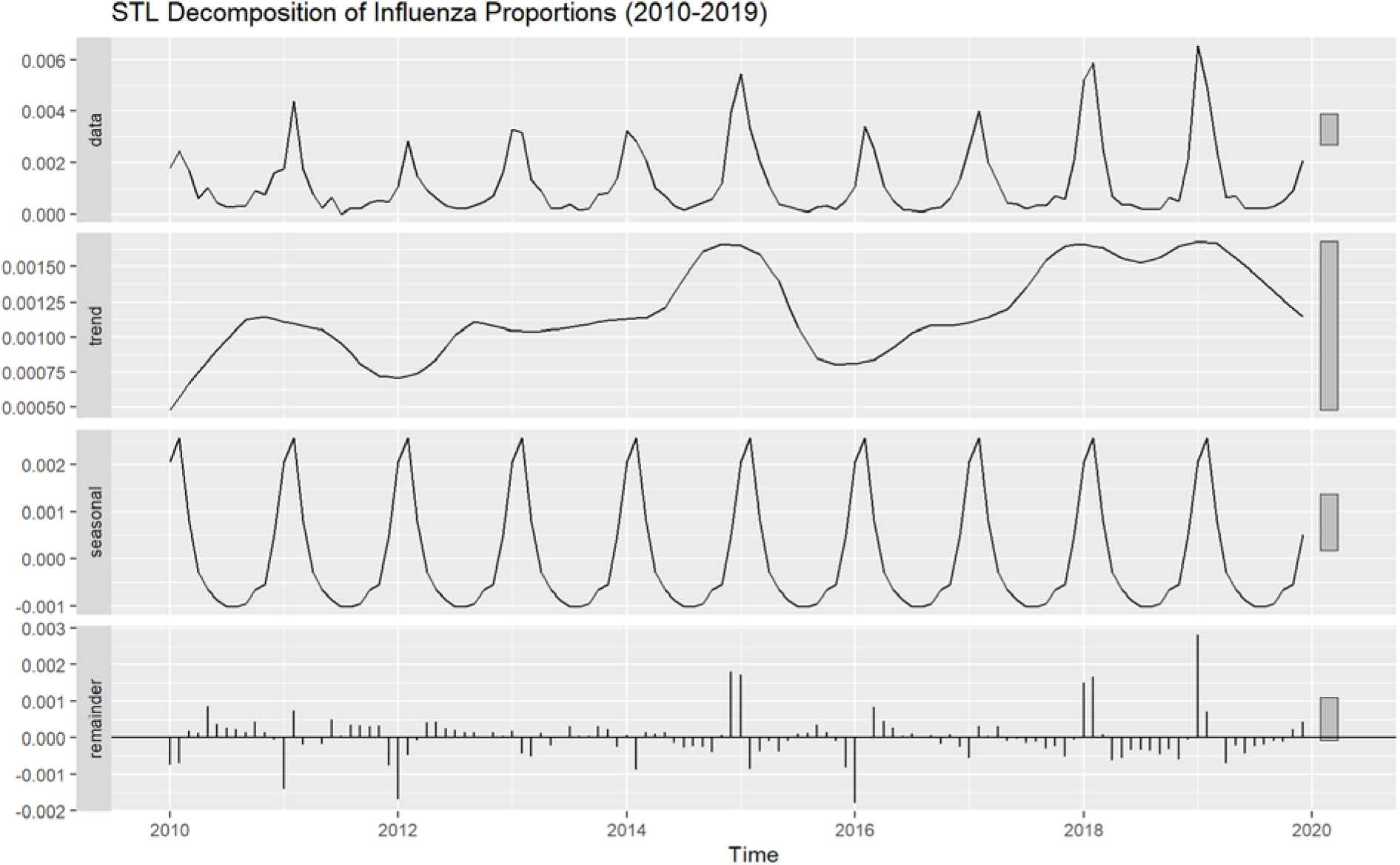
STL Decomposition of Influenza Report Proportions Over Time. The raw time series (top panel) was decomposed into a long-term trend component (second panel), a seasonal component reflecting recurring within-year fluctuations (third panel), and a remainder capturing unexplained variation (bottom panel). Strong seasonal structure is observed, consistent with a high seasonality strength of 0.8 calculated from this decomposition.

**eFigure 3.**
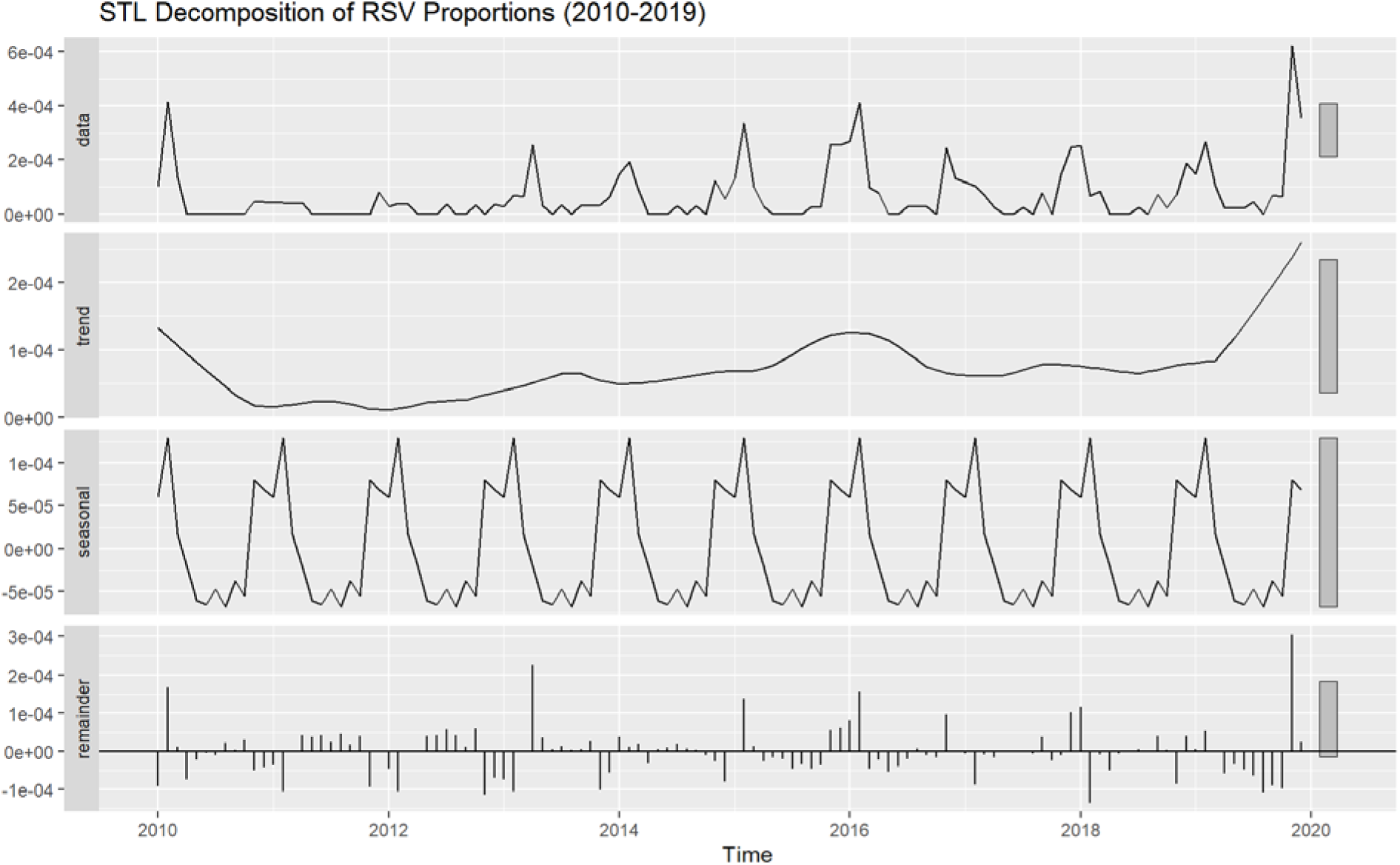
STL Decomposition of RSV Report Proportions Over Time. The raw time series (top panel) was decomposed into a long-term trend component (second panel), a seasonal component reflecting recurring within-year fluctuations (third panel), and a remainder capturing unexplained variation (bottom panel). Moderate seasonal structure is evident, consistent with a seasonality strength of 0.508 calculated from this decomposition.

**eFigure 4.**
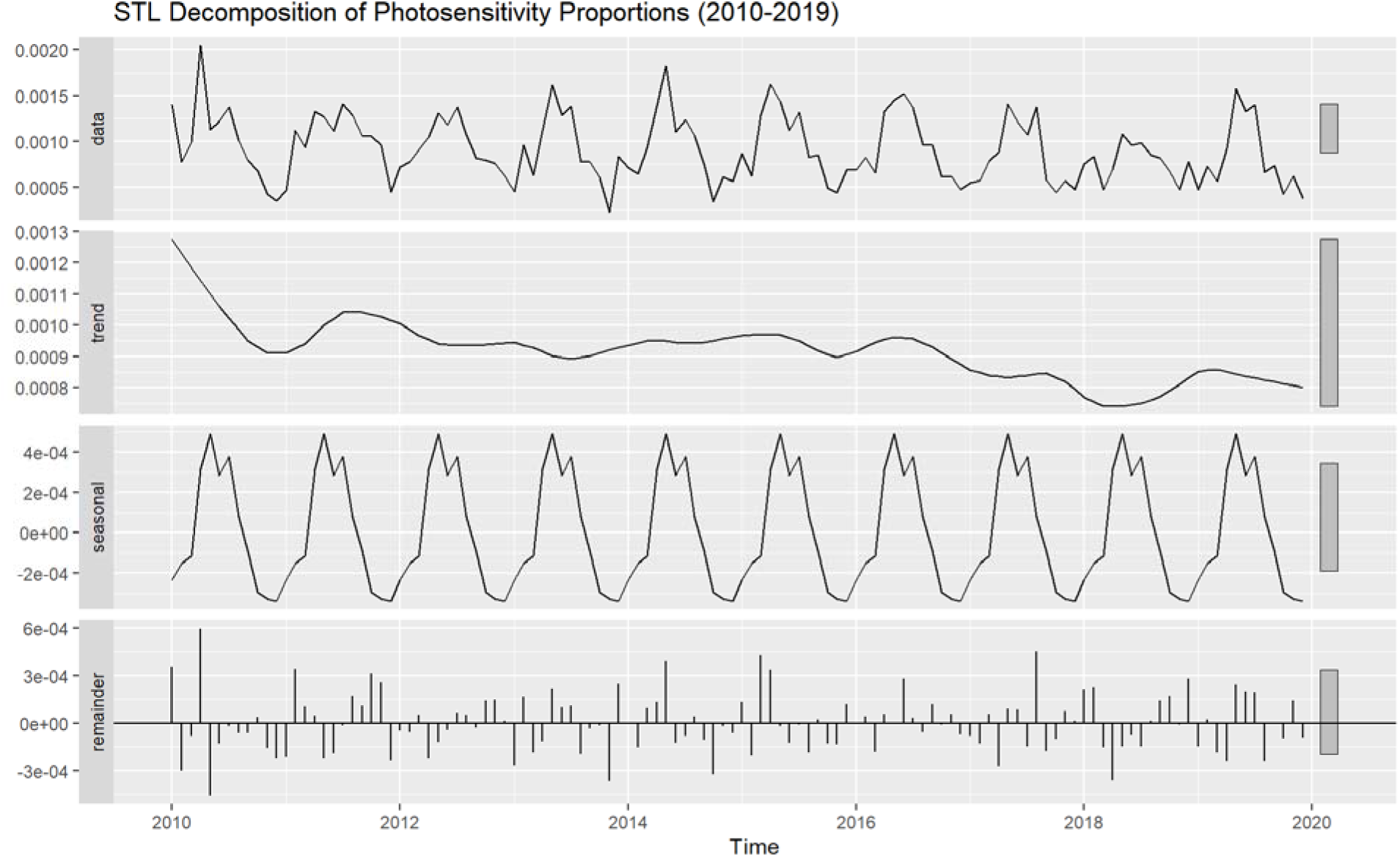
STL Decomposition of Photosensitivity Report Proportions Over Time. The raw time series (top panel) was decomposed into a long-term trend component (second panel), a seasonal component reflecting recurring within-year fluctuations (third panel), and a remainder capturing unexplained variation (bottom panel). Strong seasonal structure is evident, consistent with high seasonality strength (0.704) calculated from this decomposition.

**eFigure 5.**
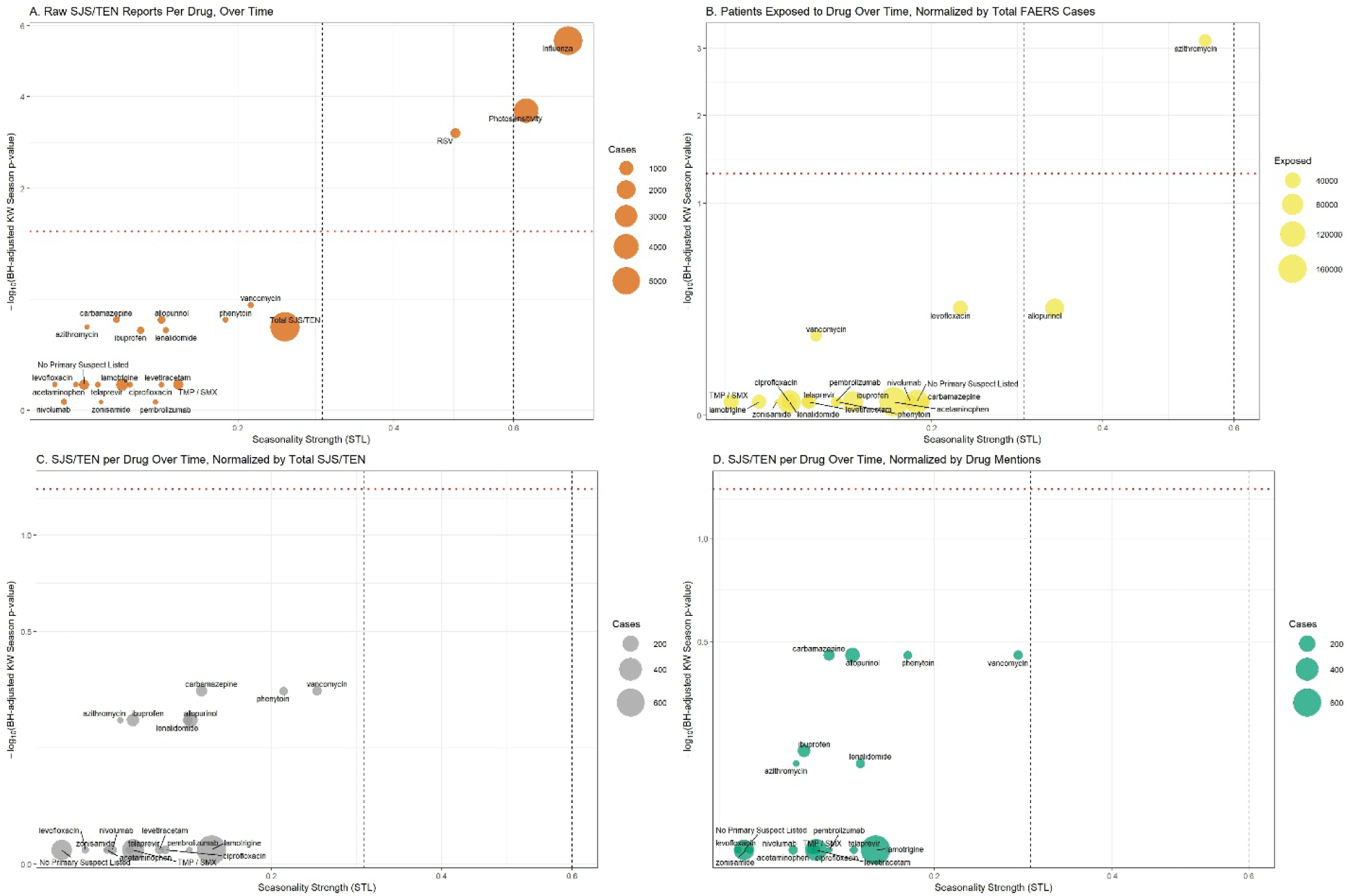
Seasonality Strength and Statistical Significance of SJS/TEN Reporting Across Drugs Using Multiple Normalization Strategies. Bubble plots showing seasonality strength (x-axis, from STL decomposition) versus statistical significance of seasonal variation (-log_10_ Benjamini-Hochberg adjusted p-value from Kruskal-Wallis tests; y-axis) for drugs associated with at least 50 SJS/TEN cases in FAERS from 2010–2019. Bubble size reflects the number of cases or exposures. Dashed vertical lines at 0.3 and 0.6 demarcate weak, moderate, and strong seasonality thresholds. The red horizontal line marks the significance threshold (adjusted p = 0.05). (A) Raw SJS/TEN reports per drug over time. (B) Number of patients exposed to each drug over time, normalized by total FAERS reports. (C) SJS/TEN reports per drug, normalized by total SJS/TEN cases. (D) SJS/TEN reports per drug, normalized by drug mentions (as a proxy for exposure).

**eFigure 6.**
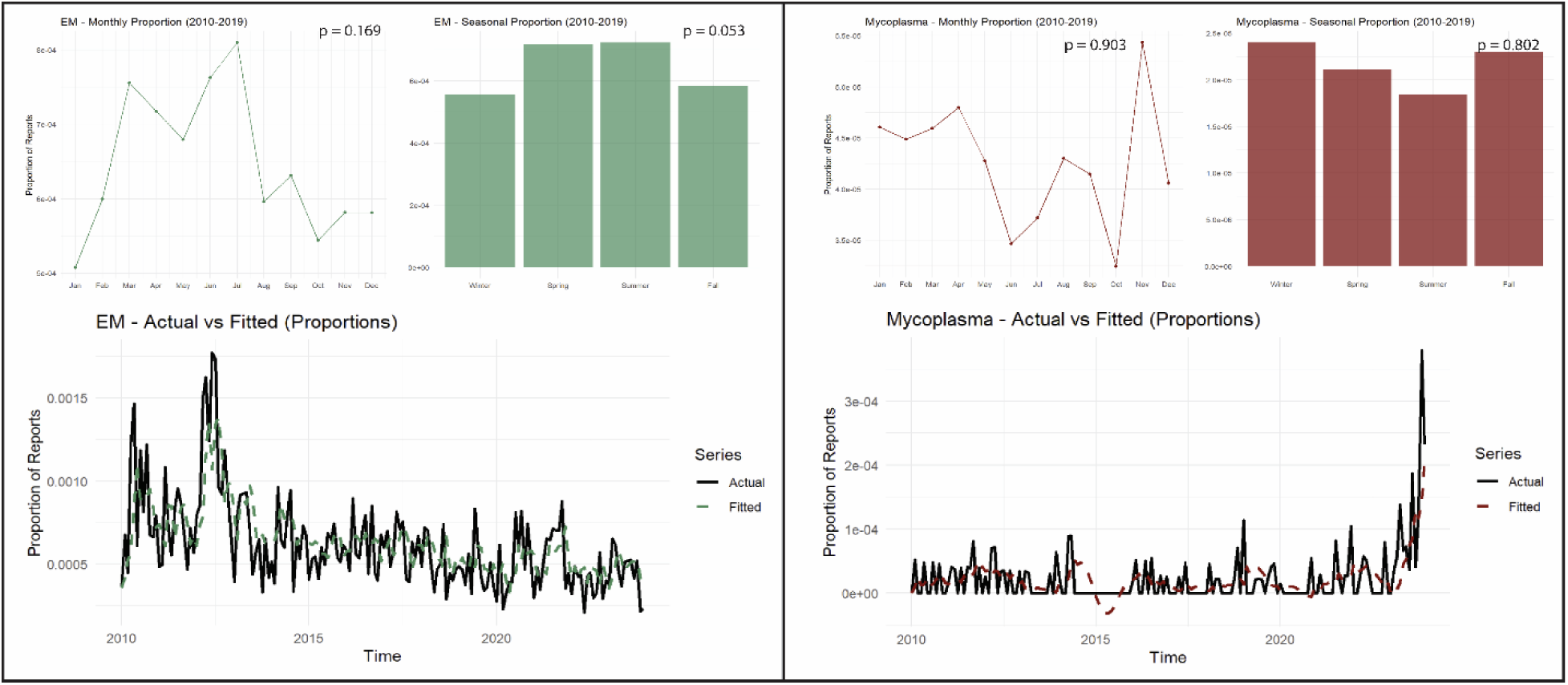
Respiratory Mycoplasma and EM Summary. Neither EM nor respiratory *Mycoplasma* infection showed statistically significant seasonality by month or by season from 2010-2019, or by STL decomposition (0.256 for EM, 0.177 for *Mycoplasma*). A marked increase in *Mycoplasma* reporting was noted near the end of the study period (2023).

**eTable 1.**
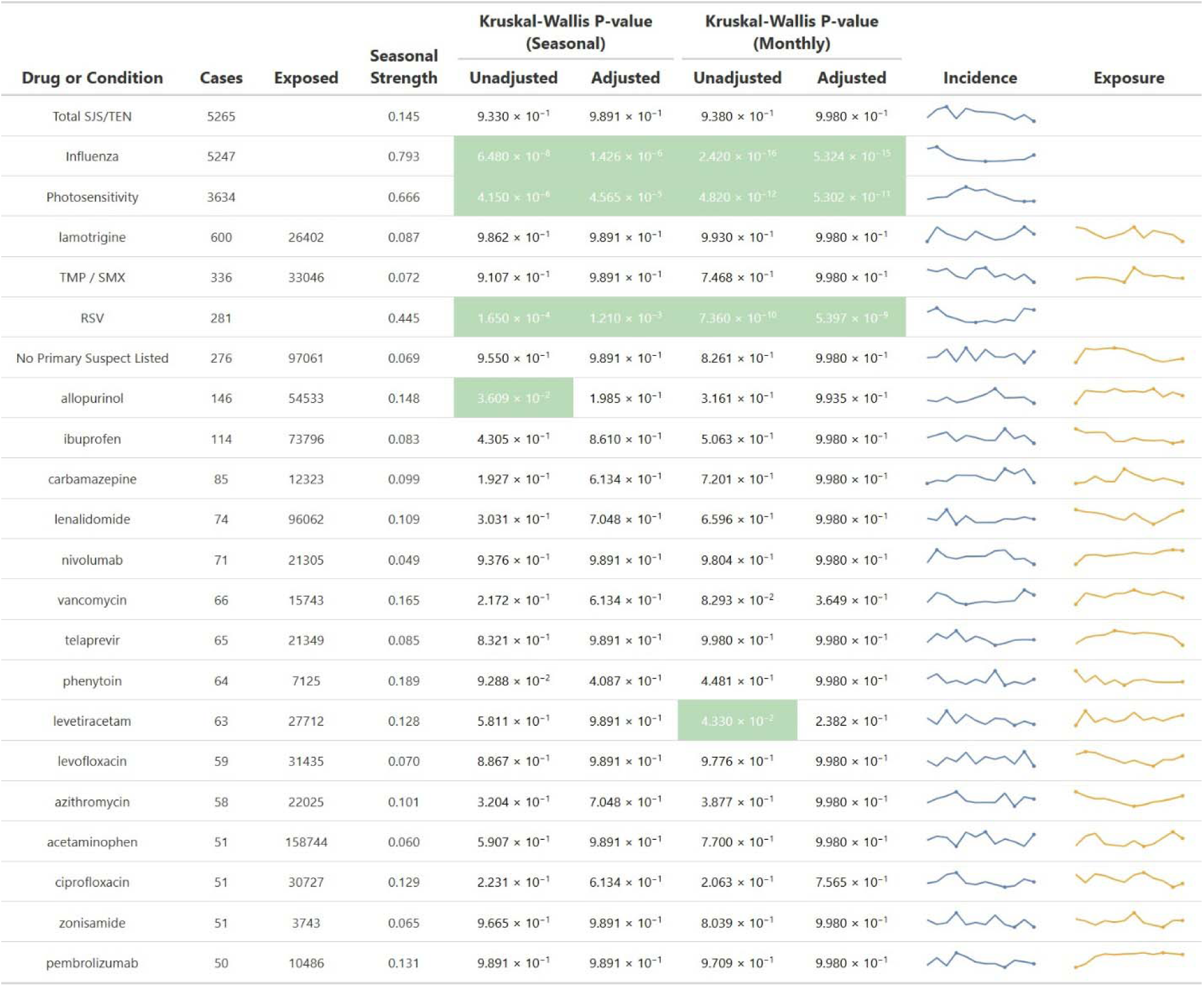
Seasonality Measures – Northern Hemisphere (Restricted)

**eTable 2.**
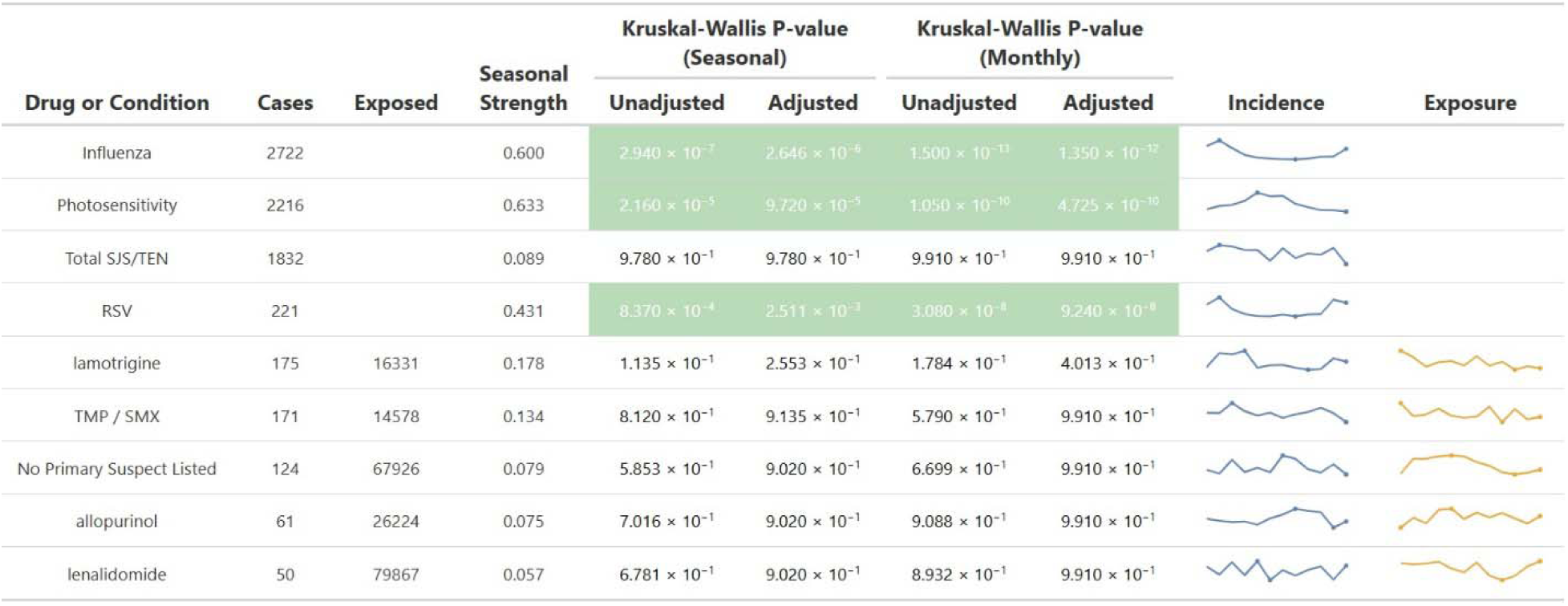
Seasonality Measures – US Only.

**eTable 3.** Time Series and Forecasting Accuracy.

**eTable 2. Seasonality Measures – US Only** Forecast Error Metrics and Model Diagnostics
| Outcome | MSE (Pre-COVID) | RMSE (Pre-COVID) | MSE (Post-COVID) | RMSE (Post-COVID) | ARIMA Model | Model Coefficients | Ljung-Box Test |
| --- | --- | --- | --- | --- | --- | --- | --- |
| SJS/TEN | 1.05e-07 | 3.24e-04 | 2.52e-07 | 5.02e-04 | ARIMA(0,1,1) with drift | ma1 = -0.871<br>drift = -1.16e-05 | Q* = 30.3<br>df = 23<br>p = 0.141 |
| Influenza | 3.53e-07 | 5.94e-04 | 2.82e-06 | 1.68e-03 | ARIMA(1,0,0)(2,1,2)[12] | ar1 = 0.489<br>sar1 = 0.698<br>sar2 = -0.479<br>sma1 = -1.42<br>sma2 = 0.759 | Q* = 14<br>df = 19<br>p = 0.782 |
| RSV | 7.17e-09 | 8.47e-05 | 3.36e-08 | 1.83e-04 | ARIMA(3,1,2) | ar1 = 1.17<br>ar2 = -0.116<br>ar3 = -0.467<br>ma1 = -1.75<br>ma2 = 0.885 | Q* = 26.9<br>df = 19<br>p = 0.107 |
| Photosensitivity | 4.50e-08 | 2.12e-04 | 4.69e-08 | 2.17e-04 | ARIMA(0,0,0)(0,1,1)[12] with drift | sma1 = -0.879<br>drift = -2.07e-06 | Q* = 23.7<br>df = 23<br>p = 0.423 |

## Notes

### Author Declarations

FDA FAERS Database (https://www.fda.gov/drugs/fdas-adverse-event-reporting-system-faers/fda-adverse-event-reporting-system-faers-public-dashboard)

