## Supplementary material for "Seasonality and Trends in Stevens-Johnson Syndrome/Toxic Epidermal Necrolysis Before and During the COVID-19 Pandemic: A Pharmacovigilance Study": eAppendix

**eAppendix. Expanded Methods**

*Geographical Classification*

| **Country Name** | **ISO 3166-1 Alpha-2 Code** | **Hemisphere** |
| --- | --- | --- |
| Afghanistan | AF | Northern |
| Albania | AL | Northern |
| Algeria | DZ | Northern |
| American Samoa | AS | Southern |
| Andorra | AD | Northern |
| Angola | AO | Southern |
| Anguilla | AI | Northern |
| Antigua and Barbuda | AG | Northern |
| Argentina | AR | Southern |
| Armenia | AM | Northern |
| Aruba | AW | Northern |
| Australia | AU | Southern |
| Austria | AT | Northern |
| Azerbaijan | AZ | Northern |
| Bahamas | BS | Northern |
| Bahrain | BH | Northern |
| Bangladesh | BD | Northern |
| Barbados | BB | Northern |
| Belarus | BY | Northern |
| Belgium | BE | Northern |
| Belize | BZ | Northern |
| Benin | BJ | Northern |
| Bermuda | BM | Northern |
| Bhutan | BT | Northern |
| Bolivia | BO | Southern |
| Bonaire, Sint Eustatius and Saba | BQ | Northern |
| Bosnia and Herzegovina | BA | Northern |
| Botswana | BW | Southern |
| Brazil | BR | Equatorial |
| British Indian Ocean Territory | IO | Southern |
| Brunei Darussalam | BN | Northern |
| Bulgaria | BG | Northern |
| Burkina Faso | BF | Northern |
| Burundi | BI | Southern |
| Cabo Verde | CV | Northern |
| Cambodia | KH | Northern |
| Cameroon | CM | Northern |
| Canada | CA | Northern |
| Cayman Islands | KY | Northern |
| Central African Republic | CF | Northern |
| Chad | TD | Northern |
| Chile | CL | Southern |
| China | CN | Northern |
| Christmas Island | CX | Southern |
| Colombia | CO | Equatorial |
| Comoros | KM | Southern |
| Congo | CG | Equatorial |
| Congo (DRC) | CD | Equatorial |
| Costa Rica | CR | Northern |
| Cote d'Ivoire | CI | Northern |
| Croatia | HR | Northern |
| Cuba | CU | Northern |
| Curacao | CW | Northern |
| Cyprus | CY | Northern |
| Czech Republic | CZ | Northern |
| Denmark | DK | Northern |
| Dominica | DM | Northern |
| Dominican Republic | DO | Northern |
| Ecuador | EC | Equatorial |
| Egypt | EG | Northern |
| El Salvador | SV | Northern |
| Eritrea | ER | Northern |
| Eswatini | SZ | Southern |
| Estonia | EE | Northern |
| Ethiopia | ET | Northern |
| Falkland Islands (Malvinas) | FK | Southern |
| Faroe Islands | FO | Northern |
| Fiji | FJ | Southern |
| Finland | FI | Northern |
| France | FR | Northern |
| French Guiana | GF | Northern |
| French Polynesia | PF | Southern |
| French Southern Territories | TF | Southern |
| Gabon | GA | Equatorial |
| Gambia | GM | Northern |
| Georgia | GE | Northern |
| Germany | DE | Northern |
| Ghana | GH | Northern |
| Gibraltar | GI | Northern |
| Greenland | GL | Northern |
| Grenada | GD | Northern |
| Guadeloupe | GP | Northern |
| Guam | GU | Northern |
| Guernsey | GG | Northern |
| Guatemala | GT | Northern |
| Guinea | GN | Northern |
| Guinea-Bissau | GW | Northern |
| Guyana | GY | Northern |
| Haiti | HT | Northern |
| Holy See (Vatican City State) | VA | Northern |
| Honduras | HN | Northern |
| Hong Kong | HK | Northern |
| Hungary | HU | Northern |
| Iceland | IS | Northern |
| India | IN | Northern |
| Indonesia | ID | Equatorial |
| Iran | IR | Northern |
| Iraq | IQ | Northern |
| Ireland | IE | Northern |
| Isle of Man | IM | Northern |
| Israel | IL | Northern |
| Italy | IT | Northern |
| Jamaica | JM | Northern |
| Japan | JP | Northern |
| Jersey | JE | Northern |
| Jordan | JO | Northern |
| Kazakhstan | KZ | Northern |
| Kenya | KE | Equatorial |
| Kiribati | KI | Equatorial |
| Kuwait | KW | Northern |
| Kyrgyzstan | KG | Northern |
| Lao People's Democratic Republic | LA | Northern |
| Latvia | LV | Northern |
| Lebanon | LB | Northern |
| Lesotho | LS | Southern |
| Liberia | LR | Northern |
| Libya | LY | Northern |
| Liechtenstein | LI | Northern |
| Lithuania | LT | Northern |
| Luxembourg | LU | Northern |
| Macao | MO | Northern |
| Madagascar | MG | Southern |
| Malawi | MW | Southern |
| Malaysia | MY | Equatorial |
| Maldives | MV | Equatorial |
| Mali | ML | Northern |
| Malta | MT | Northern |
| Martinique | MQ | Northern |
| Mauritania | MR | Northern |
| Mauritius | MU | Southern |
| Mayotte | YT | Southern |
| Mexico | MX | Northern |
| Federated States of Micronesia | FM | Northern |
| Moldova | MD | Northern |
| Monaco | MC | Northern |
| Mongolia | MN | Northern |
| Montserrat | MS | Northern |
| Montenegro | ME | Northern |
| Morocco | MA | Northern |
| Mozambique | MZ | Southern |
| Myanmar | MM | Northern |
| Nauru | NR | Southern |
| Nepal | NP | Northern |
| Netherlands | NL | Northern |
| New Caledonia | NC | Southern |
| New Zealand | NZ | Southern |
| Nicaragua | NI | Northern |
| Niger | NE | Northern |
| Nigeria | NG | Northern |
| Norfolk Island | NF | Southern |
| North Korea | KP | Northern |
| North Macedonia | MK | Northern |
| Northern Mariana Islands | MP | Northern |
| Norway | NO | Northern |
| Oman | OM | Northern |
| Pakistan | PK | Northern |
| Palau | PW | Northern |
| State of Palestine | PS | Northern |
| Panama | PA | Northern |
| Papua New Guinea | PG | Equatorial |
| Paraguay | PY | Southern |
| Peru | PE | Southern |
| Philippines | PH | Northern |
| Pitcairn | PN | Southern |
| Poland | PL | Northern |
| Portugal | PT | Northern |
| Puerto Rico | PR | Northern |
| Qatar | QA | Northern |
| Reunion | RE | Southern |
| Romania | RO | Northern |
| Russia | RU | Northern |
| Rwanda | RW | Southern |
| Saint Barthelemy | BL | Northern |
| Saint Helena, Ascension and Tristan da Cunha | SH | Southern |
| Saint Kitts and Nevis | KN | Northern |
| Saint Lucia | LC | Northern |
| Saint Martin (French part) | MF | Northern |
| Saint Pierre and Miquelon | PM | Northern |
| Saint Vincent and the Grenadines | VC | Northern |
| Samoa | WS | Southern |
| San Marino | SM | Northern |
| Sao Tome and Principe | ST | Equatorial |
| Saudi Arabia | SA | Northern |
| Senegal | SN | Northern |
| Serbia | RS | Northern |
| Seychelles | SC | Southern |
| Sierra Leone | SL | Northern |
| Singapore | SG | Northern |
| Sint Maarten (Dutch part) | SX | Northern |
| Slovakia | SK | Northern |
| Slovenia | SI | Northern |
| Solomon Islands | SB | Southern |
| Somalia | SO | Equatorial |
| South Africa | ZA | Southern |
| South Korea | KR | Northern |
| South Sudan | SS | Northern |
| Spain | ES | Northern |
| Sri Lanka | LK | Northern |
| Sudan | SD | Northern |
| Suriname | SR | Northern |
| Svalbard and Jan Mayen | SJ | Northern |
| Sweden | SE | Northern |
| Switzerland | CH | Northern |
| Syria | SY | Northern |
| Taiwan | TW | Northern |
| Tajikistan | TJ | Northern |
| Tanzania | TZ | Southern |
| Thailand | TH | Northern |
| Timor-Leste | TL | Southern |
| Togo | TG | Northern |
| Tokelau | TK | Southern |
| Tonga | TO | Southern |
| Trinidad and Tobago | TT | Northern |
| Tunisia | TN | Northern |
| Turkey | TR | Northern |
| Turks and Caicos Islands | TC | Northern |
| Tuvalu | TV | Southern |
| U.S. Virgin Islands | VI | Northern |
| Uganda | UG | Equatorial |
| Ukraine | UA | Northern |
| United Arab Emirates | AE | Northern |
| United Kingdom | GB | Northern |
| United States | US | Northern |
| Uruguay | UY | Southern |
| Uzbekistan | UZ | Northern |
| Vanuatu | VU | Southern |
| Venezuela | VE | Northern |
| Vietnam | VN | Northern |
| Virgin Islands, British | VG | Northern |
| Wallis and Futuna | WF | Southern |
| Western Sahara | EH | Northern |
| Yemen | YE | Northern |
| Zambia | ZM | Southern |
| Zimbabwe | ZW | Southern |
| Aland Islands | AX | Northern |

For the restricted northern hemisphere sample, the following countries were used: US, Canada, Germany, Great Britain, France, Italy, Spain, Japan, China, South Korea

**eAppendix 2. MedDRA Terms/Case Selection**

| **Condition** | **Matching Outcomes** | **Matching Indications** |
| --- | --- | --- |
| SJS/TEN | “Stevens-Johnson syndrome”, “SJS-TEN overlap”, “Toxic epidermal necrolysis” |  |
| RSV |  | “Respiratory syncytial virus infection”, “Pneumonia respiratory syncytial viral”, “Respiratory syncytial virus test positive”, “Respiratory syncytial virus bronchiolitis”, “Respiratory syncytial virus bronchitis” |
| Influenza | "Inﬂuenza", "H1N1 Inﬂuenza", "Pneumonia Inﬂuenzal", "Avian Inﬂuenza", "Inﬂuenza B Virus Test Positive", "Inﬂuenza Virus Test Positive", "Inﬂuenza A Virus Test Positive", "H3N2 Inﬂuenza", "Swine Inﬂuenza", "Inﬂuenza A Virus Infection", "Flu", "Inﬂuenza Serology Positive" |  |
| Photosensitivity | “Photosensitivity reaction”, “photosensitivity allergic reaction”, “photodermatosis” | “Photosensitivity reaction”, “Injection site photosensitivity reaction”, “Application site photosensitivity reaction”, “Administration site photosensitivity reaction”, “Infusion site photosensitivity reaction”, “Photosensitive rash”, “Photosensitivity allergic reaction” |
| Erythema Multiforme | “Erythema Multiforme” | “Erythema Multiforme” |
| *Mycoplasma* |  | “Mycoplasma Infection”, “Pneumonia Mycoplasmal”, “Bronchitis Mycoplasmal”, “Pharyngitis Mycoplasmal”, “Mycoplasma Pneumoniae Pneumonia”, “Tracheobronchitis Mycoplasmal” |

**eAppendix 3. Time Series Analysis**

*STL Decomposition*

To evaluate the presence and strength of seasonal patterns in the time series data, we employed Seasonal-Trend Decomposition using Loess (STL), implemented via the stl() function in the base R stats package. STL is a robust and flexible method that decomposes a time series into three distinct components: the **seasonal component (S)**, representing within-year recurring patterns; the **trend component (T)**, capturing long-term changes in level or slope; and the **remainder (R)**, representing residual variability not explained by either seasonality or trend.

We applied STL decomposition to the monthly time series of case proportions for each condition of interest (SJS/TEN, photosensitivity, influenza, RSV). The decomposition was performed additively, meaning that the observed time series was assumed to be the sum of the seasonal, trend, and remainder components. For the seasonal window parameter, we specified the "periodic" setting, which enforces a fixed seasonal pattern repeating identically each year. The trend window was set to its default, allowing for adaptive smoothing of long-term trends without overfitting.

To quantify the strength of seasonality in each time series, we calculated a **seasonality strength metric**, defined as the proportion of variance explained by the seasonal component relative to the combined variance of the seasonal and remainder components.

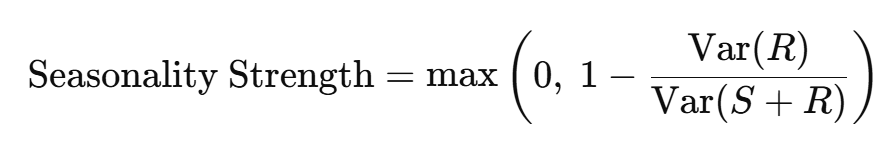

where **Var(S)** is the variance of the seasonal component and **Var(R)** is the variance of the remainder component. This metric ensures that negative values are set to zero, preventing interpretational artifacts. A value closer to 1 indicates that the seasonal component accounts for a substantial portion of the variability in the time series, reflecting strong and consistent seasonality. Conversely, values near 0 indicate minimal or negligible seasonal patterning, where most of the variability is attributed to noise or irregular fluctuations rather than consistent seasonal effects.

This quantitative approach allowed us to directly compare seasonality strength across conditions, and to contextualize findings within the broader landscape of pharmacovigilance data, where environmental and temporal influences may differentially impact various adverse events.

*SARIMA Time Series Modeling*

To model temporal trends and forecast adverse event reporting beyond the pre-pandemic period, we applied Seasonal Autoregressive Integrated Moving Average (SARIMA) models to the monthly time series of case proportions for each condition of interest. SARIMA models extend the traditional ARIMA framework by incorporating parameters that account for both non-seasonal and seasonal patterns in time series data. A SARIMA model is generally denoted as **ARIMA(p, d, q)(P, D, Q)[s]**, where:

- **p**: number of non-seasonal autoregressive (AR) terms
- **d**: number of non-seasonal differences required to achieve stationarity
- **q**: number of non-seasonal moving average (MA) terms
- **P**: number of seasonal autoregressive terms
- **D**: number of seasonal differences
- **Q**: number of seasonal moving average terms
- **s**: the length of the seasonal period (e.g., 12 for monthly data with annual seasonality)

We fit SARIMA models using the auto.arima() function from the forecast package in R, which automates the selection of optimal model parameters based on the Akaike Information Criterion (AIC), balancing model complexity with goodness-of-fit.

After model selection, we evaluated the goodness-of-fit using residual diagnostics, including the Ljung-Box test to assess for any significant autocorrelation in the residuals. Only models with non-significant Ljung-Box p-values (p > 0.05) were retained, indicating adequate capture of serial dependence. All model coefficients and fit tests are reported in **eTable 3**

We then generated forecasts for the post-COVID period (April 2020 to December 2023), comparing observed values to forecasted estimates with 95% confidence intervals. Forecast accuracy was quantified using mean squared error (MSE) and root mean squared error (RMSE), providing a metric of prediction deviation in the post-pandemic period relative to pre-pandemic trends.
